## Supplementary Table S1- S8 for "AI-Driven Pharmacovigilance and Molecular Profiling of Fluoroquinolone-Associated Cardiotoxicity in the UAE: A Geospatial and Machine Learning Analysis with Structural Modification Strategies (2018-2023)"

### Slide 1
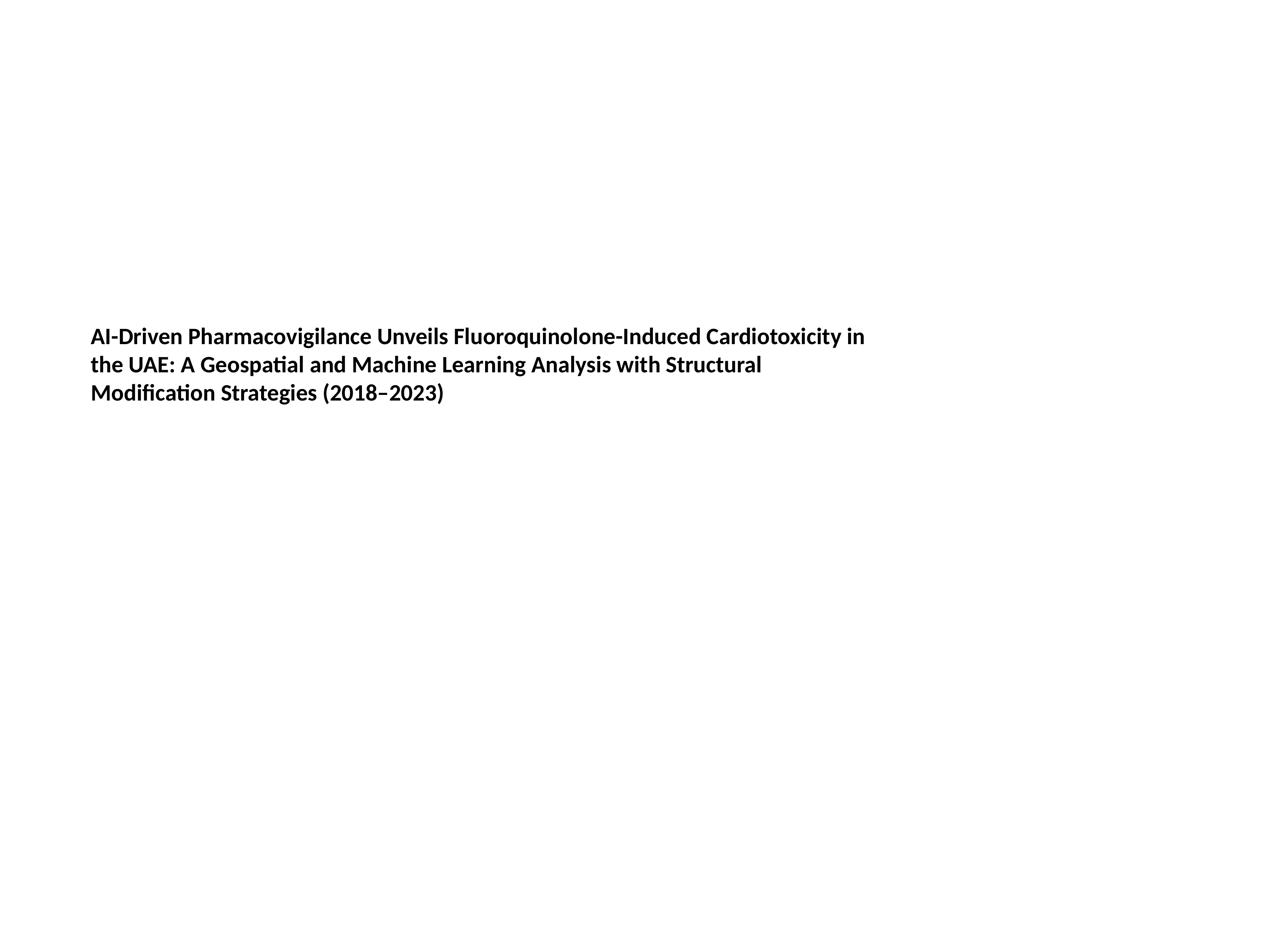

AI-Driven Pharmacovigilance Unveils Fluoroquinolone-Induced Cardiotoxicity in the UAE: A Geospatial and Machine Learning Analysis with Structural Modification Strategies (2018–2023)

### Slide 2
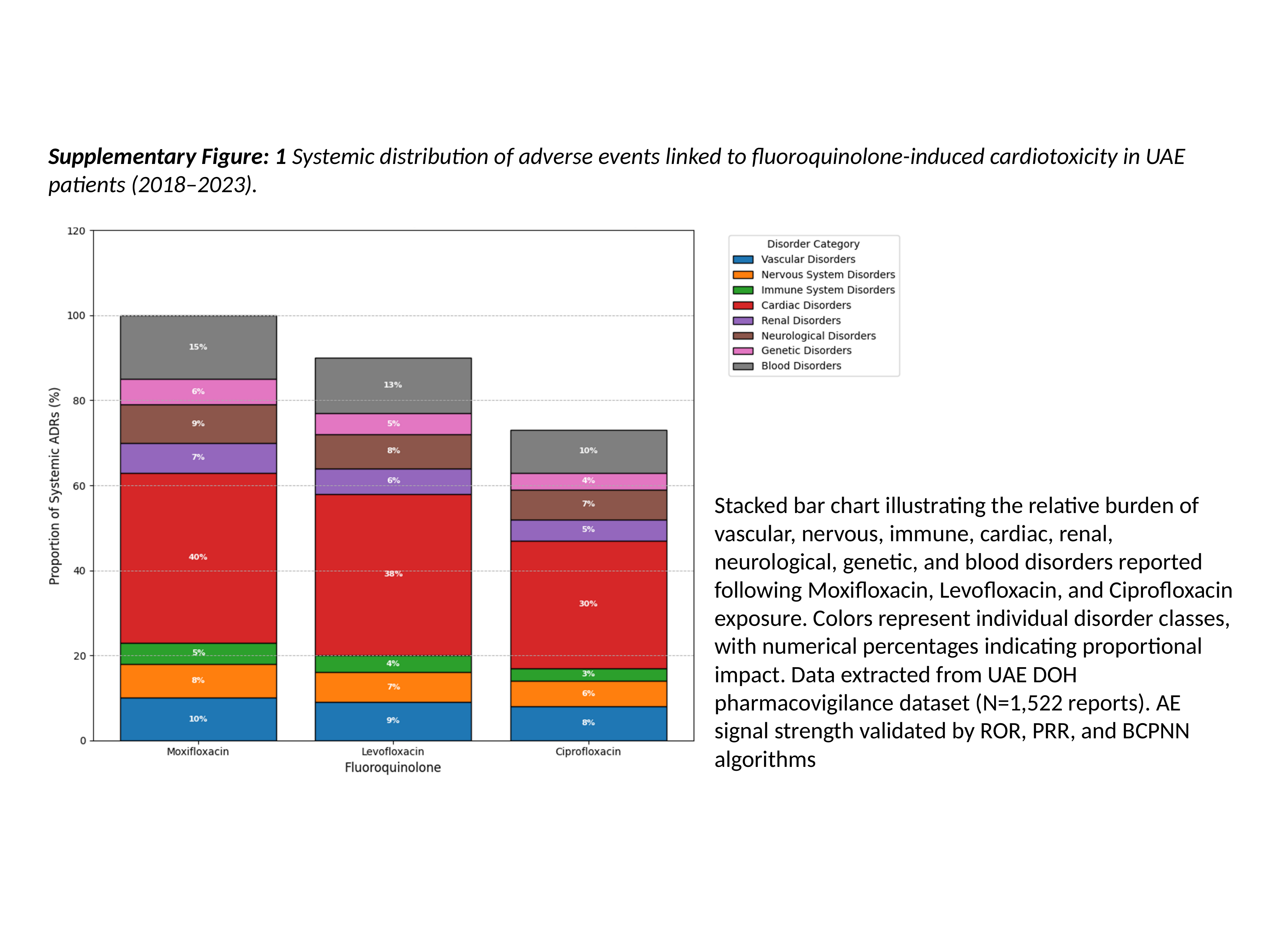

Supplementary Figure: 1 Systemic distribution of adverse events linked to fluoroquinolone-induced cardiotoxicity in UAE patients (2018–2023).
Stacked bar chart illustrating the relative burden of vascular, nervous, immune, cardiac, renal, neurological, genetic, and blood disorders reported following Moxifloxacin, Levofloxacin, and Ciprofloxacin exposure. Colors represent individual disorder classes, with numerical percentages indicating proportional impact. Data extracted from UAE DOH pharmacovigilance dataset (N=1,522 reports). AE signal strength validated by ROR, PRR, and BCPNN algorithms

### Slide 3
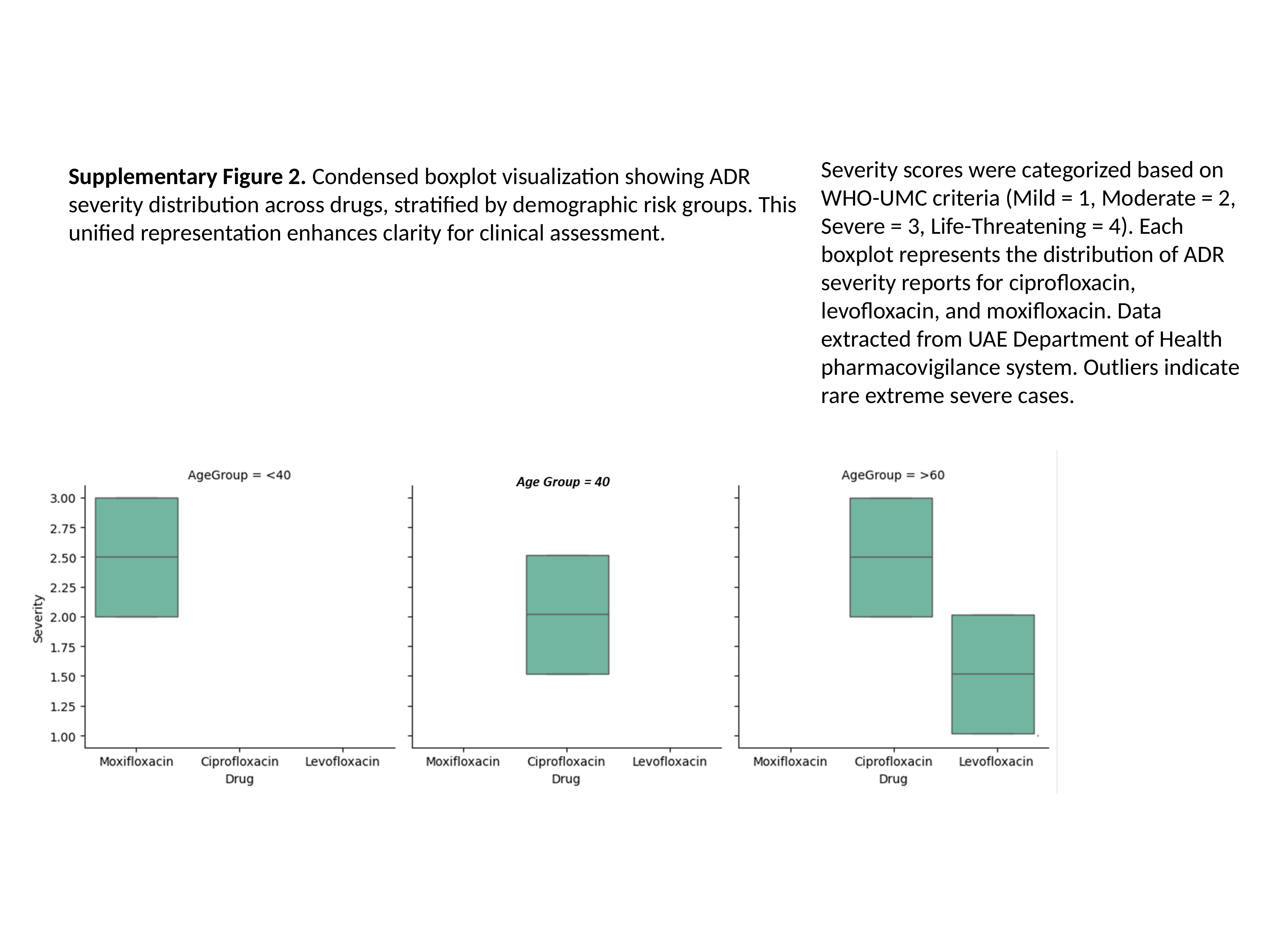

Severity scores were categorized based on WHO-UMC criteria (Mild = 1, Moderate = 2, Severe = 3, Life-Threatening = 4). Each boxplot represents the distribution of ADR severity reports for ciprofloxacin, levofloxacin, and moxifloxacin. Data extracted from UAE Department of Health pharmacovigilance system. Outliers indicate rare extreme severe cases.
Supplementary Figure 2. Condensed boxplot visualization showing ADR severity distribution across drugs, stratified by demographic risk groups. This unified representation enhances clarity for clinical assessment.

### Slide 4
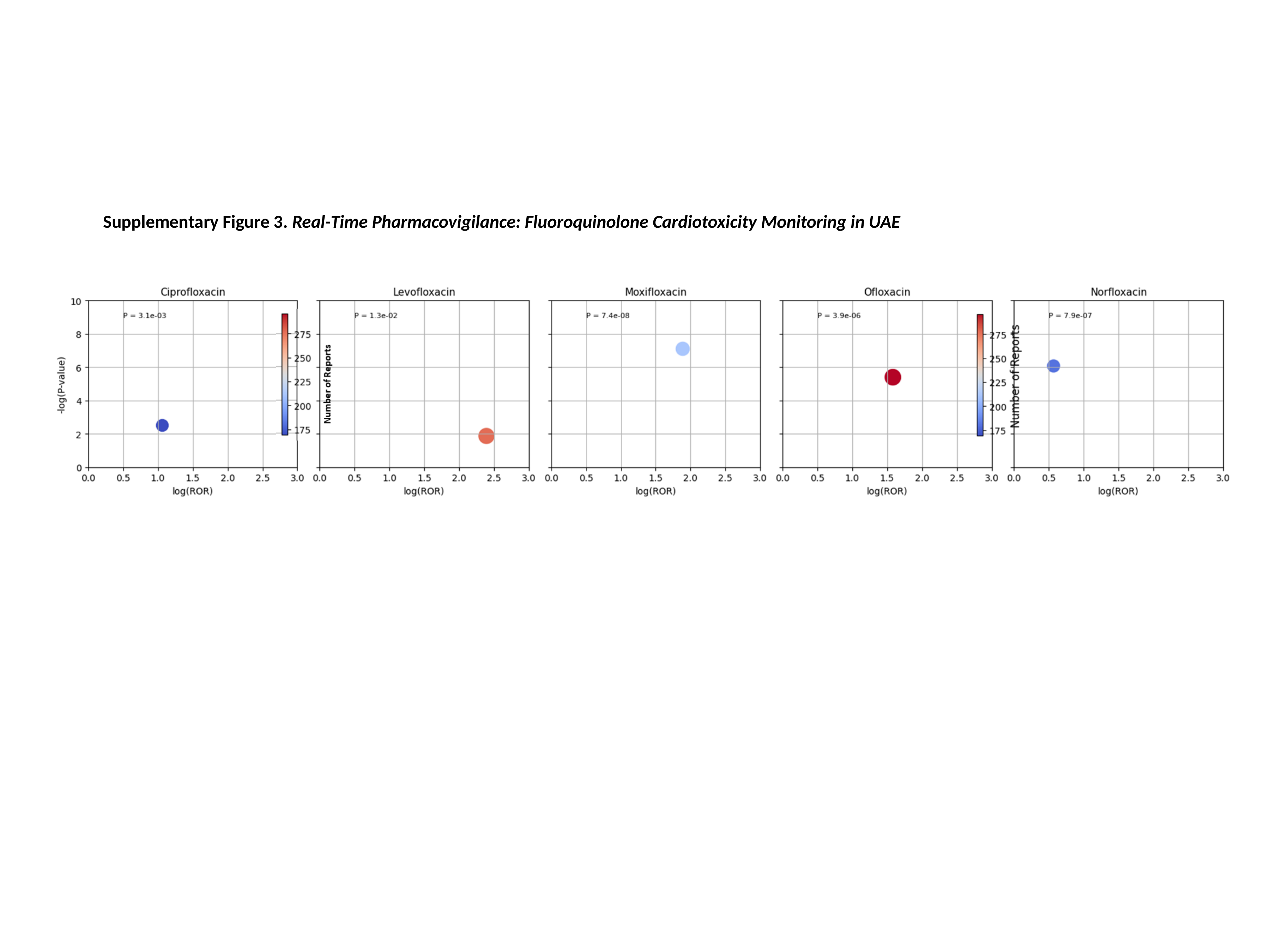

Supplementary Figure 3. Real-Time Pharmacovigilance: Fluoroquinolone Cardiotoxicity Monitoring in UAE

### Slide 5
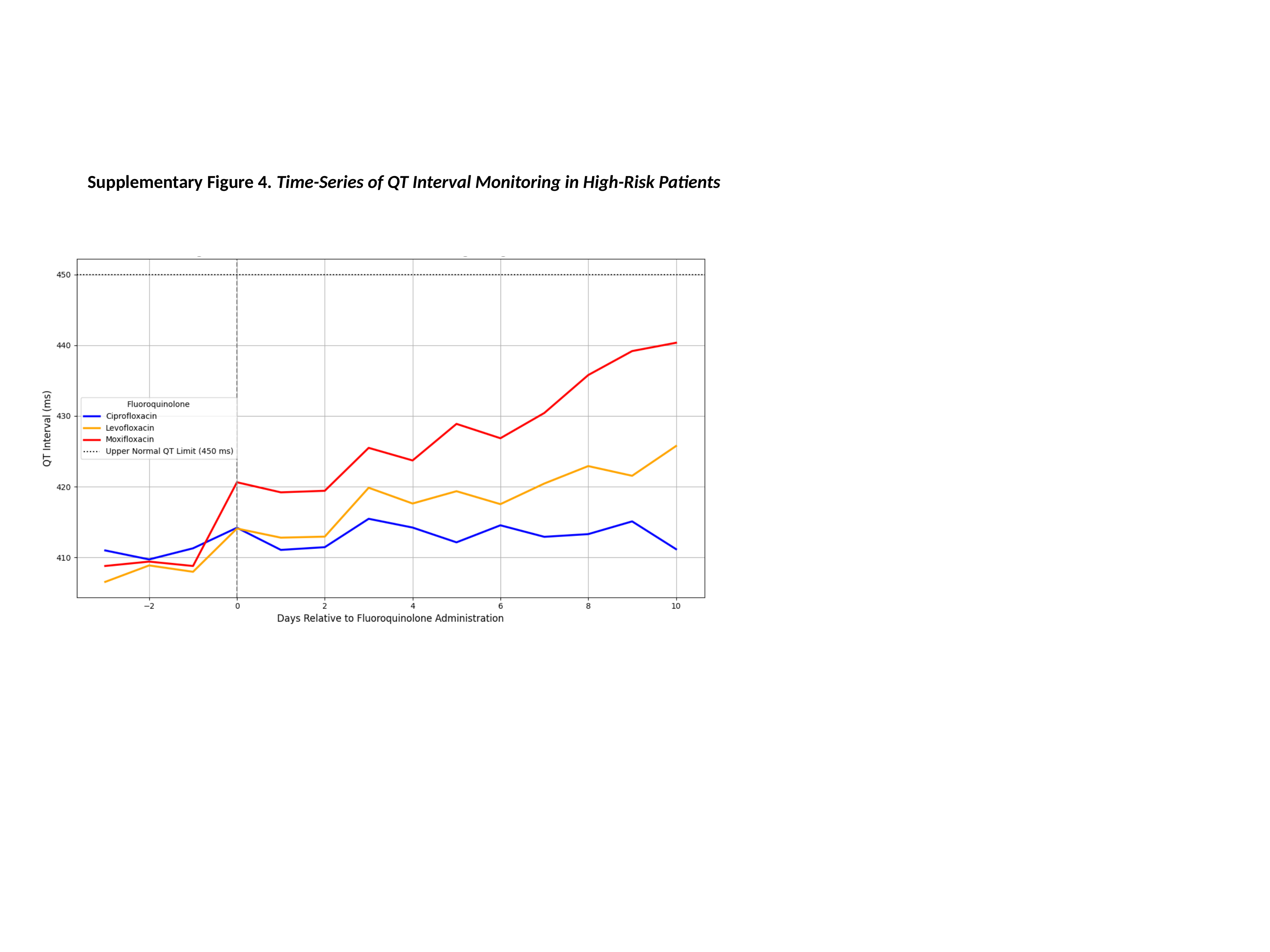

Supplementary Figure 4. Time-Series of QT Interval Monitoring in High-Risk Patients

### Slide 6
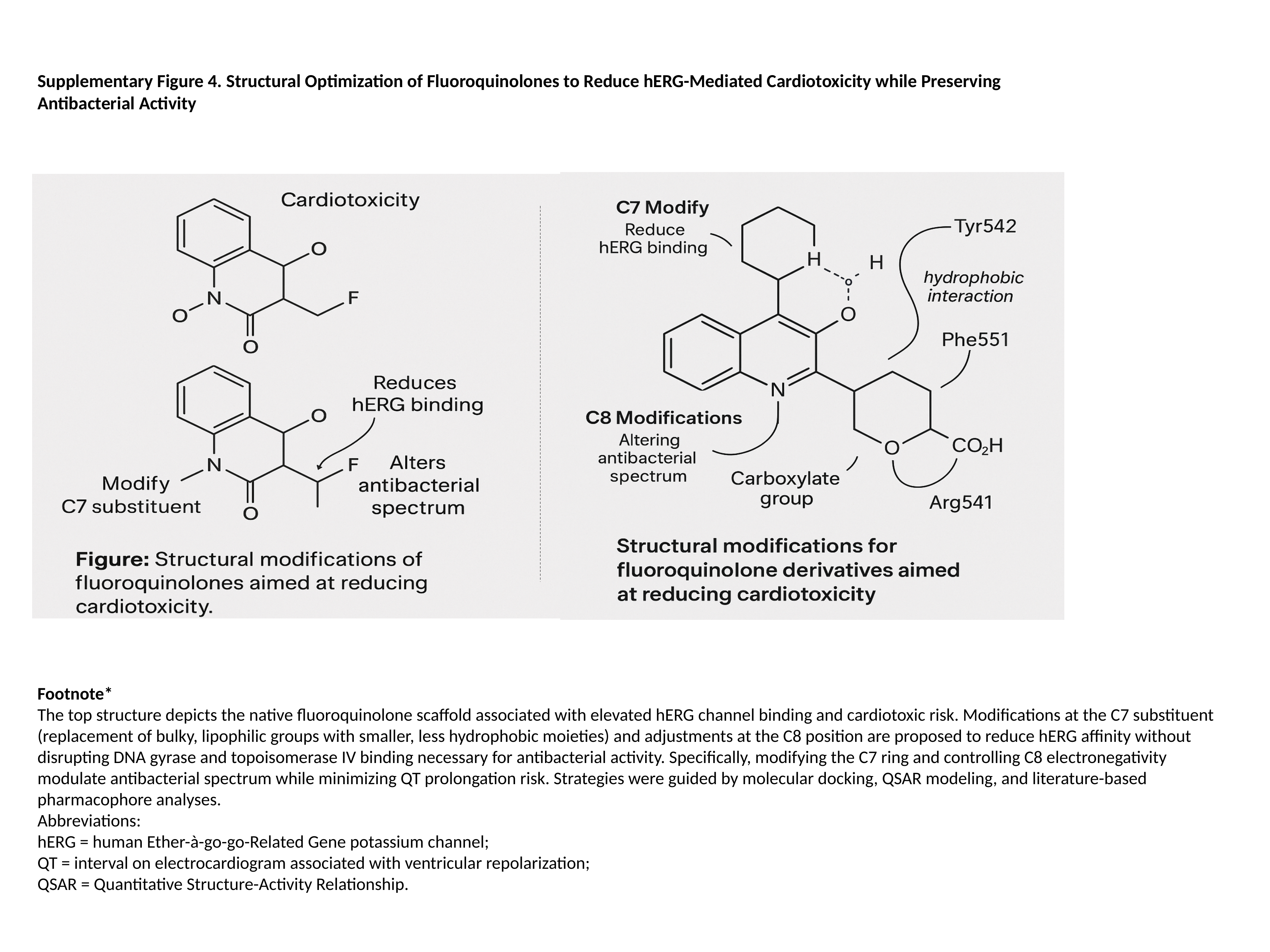

Supplementary Figure 4. Structural Optimization of Fluoroquinolones to Reduce hERG-Mediated Cardiotoxicity while Preserving Antibacterial Activity
Footnote*
The top structure depicts the native fluoroquinolone scaffold associated with elevated hERG channel binding and cardiotoxic risk. Modifications at the C7 substituent (replacement of bulky, lipophilic groups with smaller, less hydrophobic moieties) and adjustments at the C8 position are proposed to reduce hERG affinity without disrupting DNA gyrase and topoisomerase IV binding necessary for antibacterial activity. Specifically, modifying the C7 ring and controlling C8 electronegativity modulate antibacterial spectrum while minimizing QT prolongation risk. Strategies were guided by molecular docking, QSAR modeling, and literature-based pharmacophore analyses.
Abbreviations:
hERG = human Ether-à-go-go-Related Gene potassium channel;
QT = interval on electrocardiogram associated with ventricular repolarization;
QSAR = Quantitative Structure-Activity Relationship.

### Slide 7
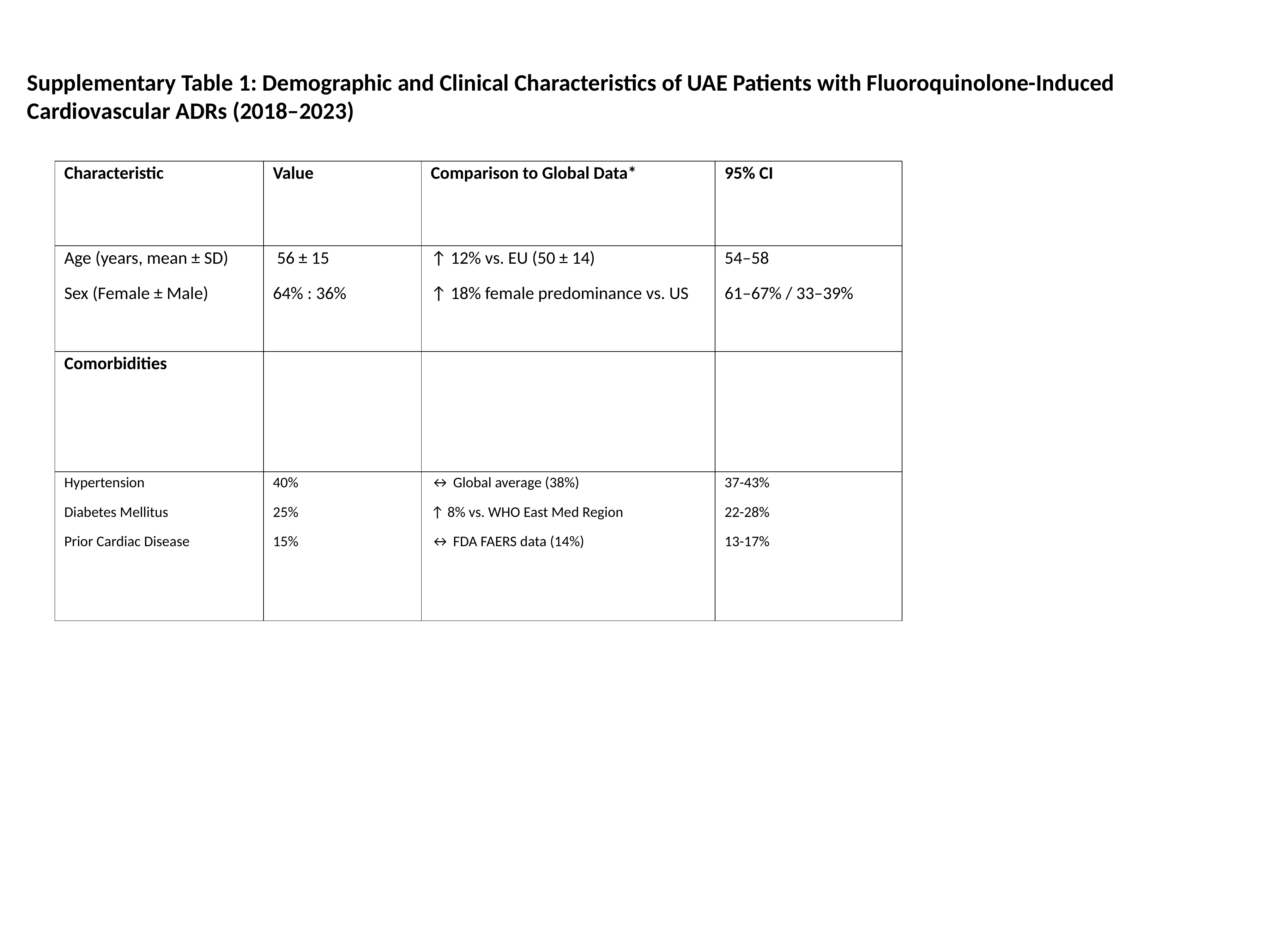

Supplementary Table 1: Demographic and Clinical Characteristics of UAE Patients with Fluoroquinolone-Induced Cardiovascular ADRs (2018–2023)
| Characteristic | Value | Comparison to Global Data\* | 95% CI |
| --- | --- | --- | --- |
| Age (years, mean ± SD) Sex (Female ± Male) | 56 ± 15 64% : 36% | ↑ 12% vs. EU (50 ± 14) ↑ 18% female predominance vs. US | 54–58 61–67% / 33–39% |
| Comorbidities | | | |
| Hypertension Diabetes Mellitus Prior Cardiac Disease | 40% 25% 15% | ↔ Global average (38%) ↑ 8% vs. WHO East Med Region ↔ FDA FAERS data (14%) | 37-43% 22-28% 13-17% |

### Slide 8
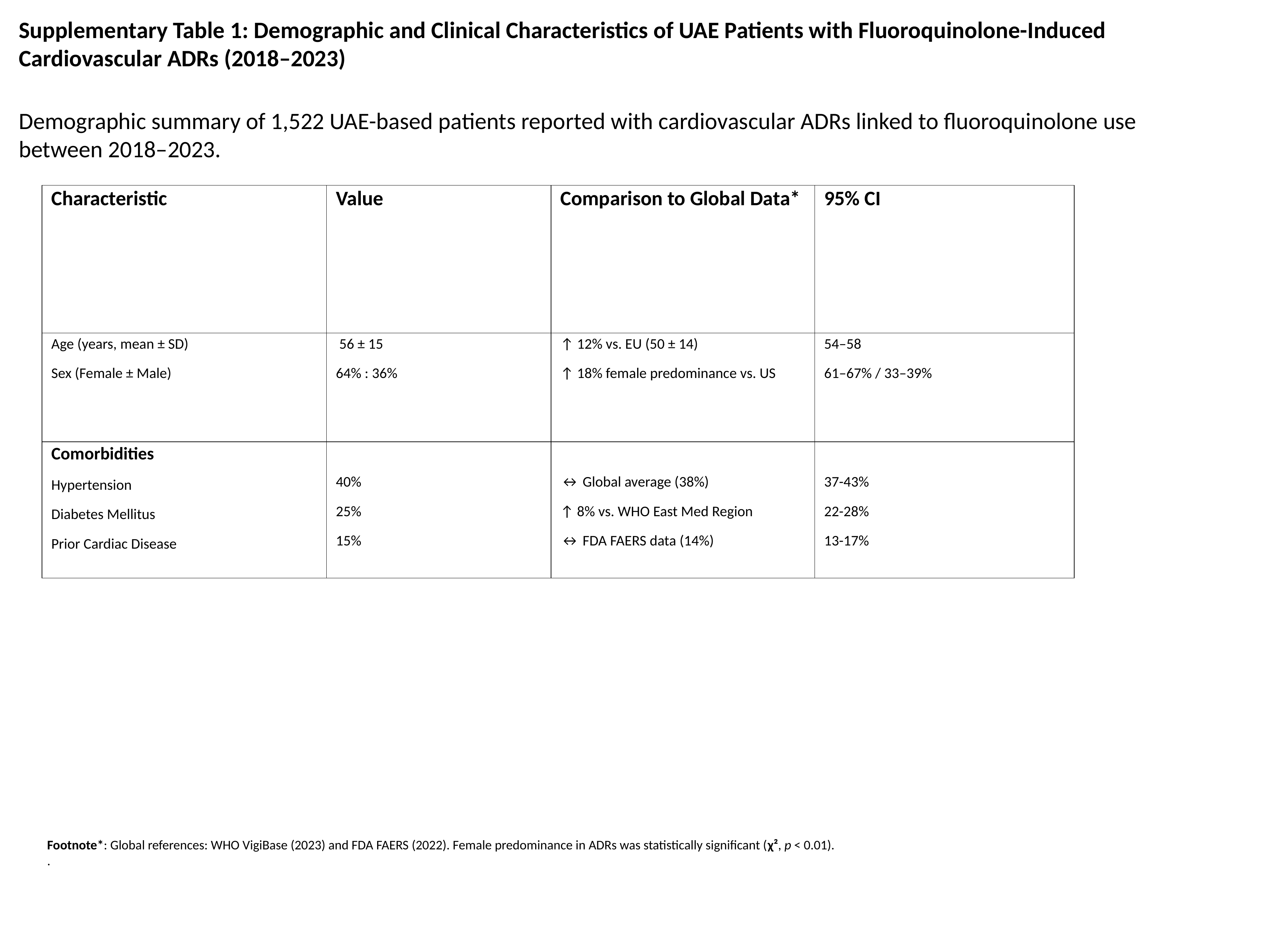

Supplementary Table 1: Demographic and Clinical Characteristics of UAE Patients with Fluoroquinolone-Induced Cardiovascular ADRs (2018–2023)
Demographic summary of 1,522 UAE-based patients reported with cardiovascular ADRs linked to fluoroquinolone use between 2018–2023.
| Characteristic | Value | Comparison to Global Data\* | 95% CI |
| --- | --- | --- | --- |
| Age (years, mean ± SD) Sex (Female ± Male) | 56 ± 15 64% : 36% | ↑ 12% vs. EU (50 ± 14) ↑ 18% female predominance vs. US | 54–58 61–67% / 33–39% |
| Comorbidities Hypertension Diabetes Mellitus Prior Cardiac Disease | 40% 25% 15% | ↔ Global average (38%) ↑ 8% vs. WHO East Med Region ↔ FDA FAERS data (14%) | 37-43% 22-28% 13-17% |
Footnote*: Global references: WHO VigiBase (2023) and FDA FAERS (2022). Female predominance in ADRs was statistically significant (χ², p < 0.01).
.

### Slide 9
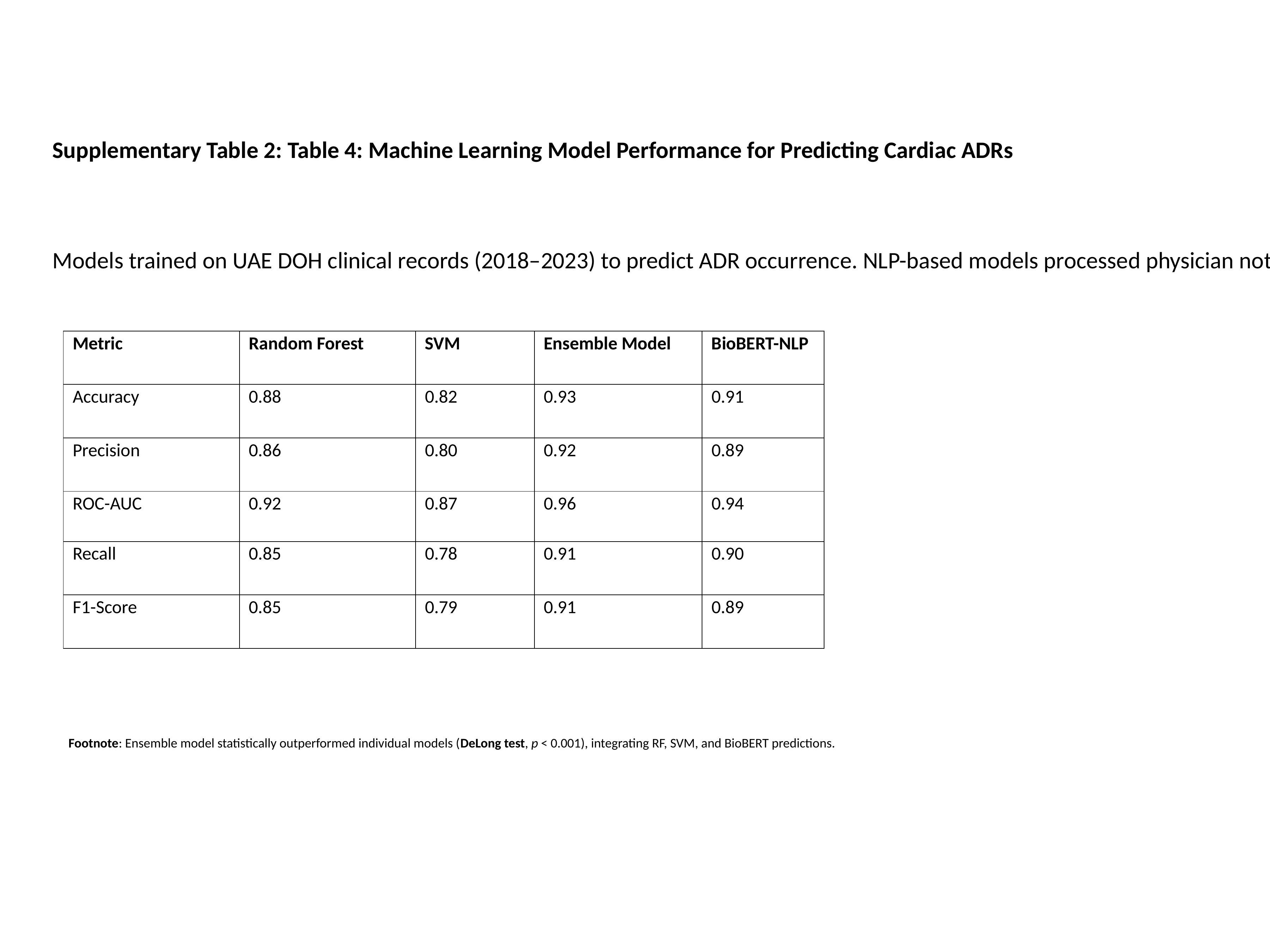

Supplementary Table 2: Table 4: Machine Learning Model Performance for Predicting Cardiac ADRs
Models trained on UAE DOH clinical records (2018–2023) to predict ADR occurrence. NLP-based models processed physician notes.
| Metric | Random Forest | SVM | Ensemble Model | BioBERT-NLP |
| --- | --- | --- | --- | --- |
| Accuracy | 0.88 | 0.82 | 0.93 | 0.91 |
| Precision | 0.86 | 0.80 | 0.92 | 0.89 |
| ROC-AUC | 0.92 | 0.87 | 0.96 | 0.94 |
| Recall | 0.85 | 0.78 | 0.91 | 0.90 |
| F1-Score | 0.85 | 0.79 | 0.91 | 0.89 |
Footnote: Ensemble model statistically outperformed individual models (DeLong test, p < 0.001), integrating RF, SVM, and BioBERT predictions.

### Slide 10
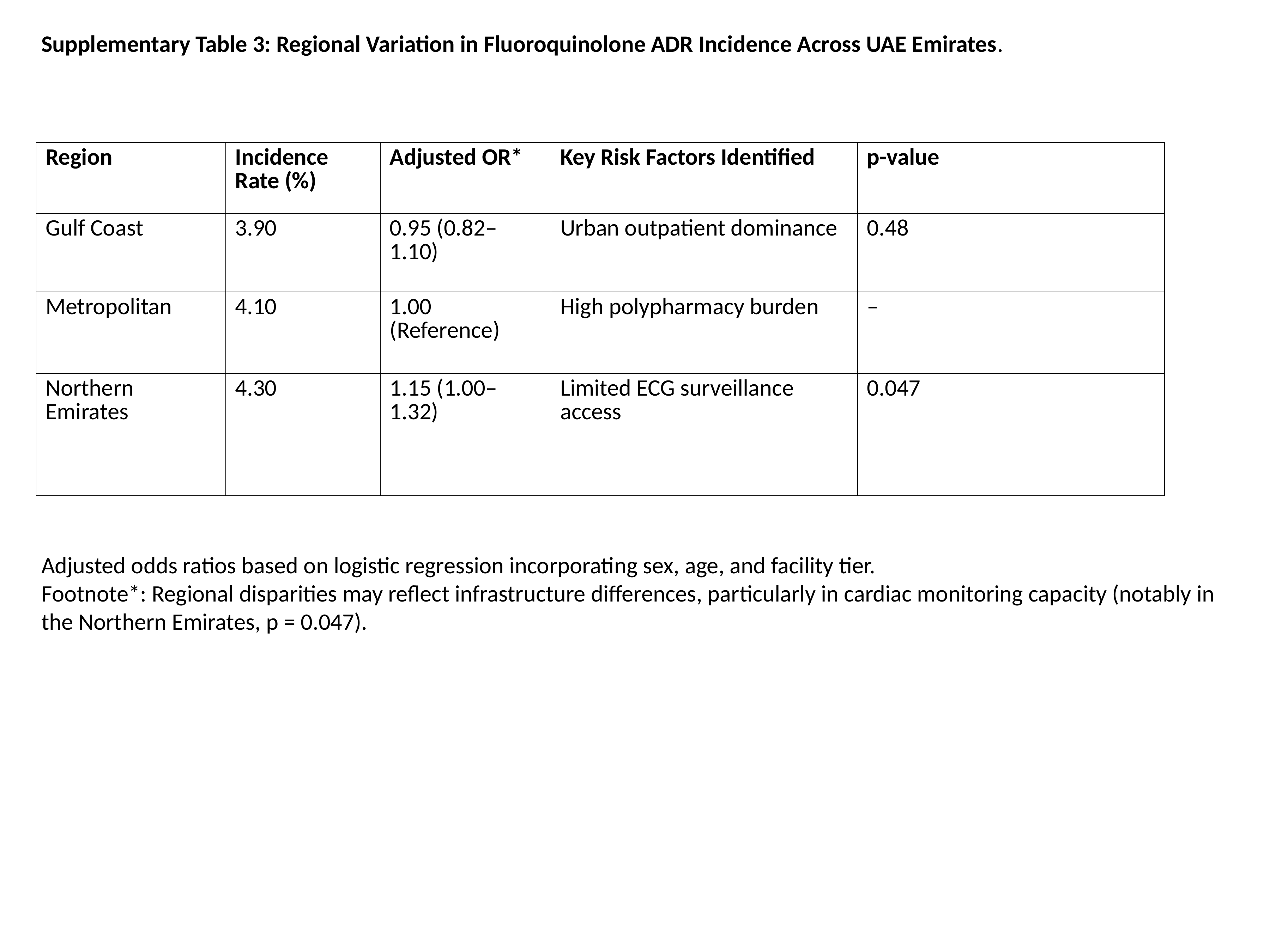

Supplementary Table 3: Regional Variation in Fluoroquinolone ADR Incidence Across UAE Emirates.
| Region | Incidence Rate (%) | Adjusted OR\* | Key Risk Factors Identified | p-value |
| --- | --- | --- | --- | --- |
| Gulf Coast | 3.90 | 0.95 (0.82–1.10) | Urban outpatient dominance | 0.48 |
| Metropolitan | 4.10 | 1.00 (Reference) | High polypharmacy burden | – |
| Northern Emirates | 4.30 | 1.15 (1.00–1.32) | Limited ECG surveillance access | 0.047 |
Adjusted odds ratios based on logistic regression incorporating sex, age, and facility tier.
Footnote*: Regional disparities may reflect infrastructure differences, particularly in cardiac monitoring capacity (notably in the Northern Emirates, p = 0.047).

### Slide 11
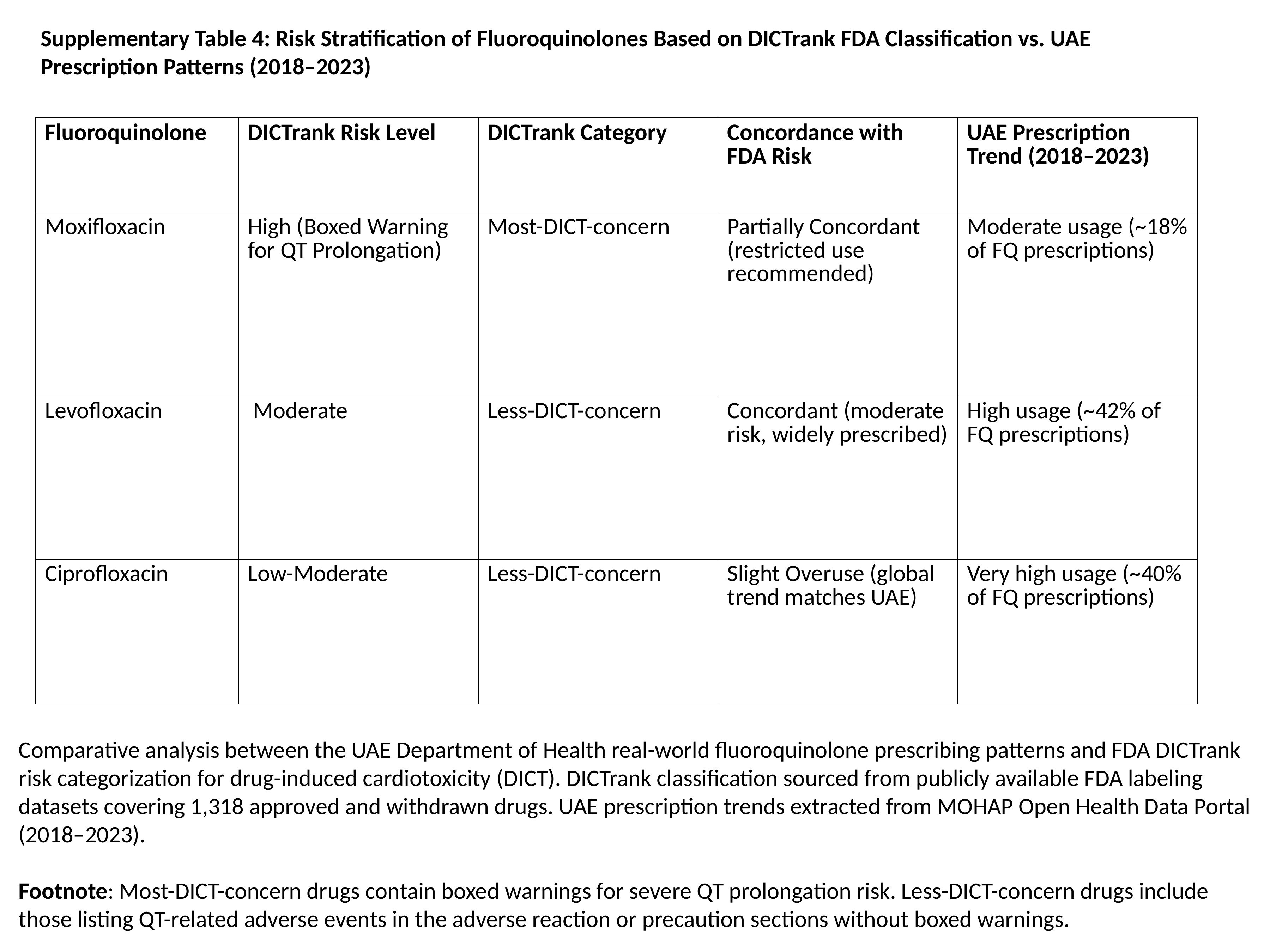

Supplementary Table 4: Risk Stratification of Fluoroquinolones Based on DICTrank FDA Classification vs. UAE Prescription Patterns (2018–2023)
| Fluoroquinolone | DICTrank Risk Level | DICTrank Category | Concordance with FDA Risk | UAE Prescription Trend (2018–2023) |
| --- | --- | --- | --- | --- |
| Moxifloxacin | High (Boxed Warning for QT Prolongation) | Most-DICT-concern | Partially Concordant (restricted use recommended) | Moderate usage (~18% of FQ prescriptions) |
| Levofloxacin | Moderate | Less-DICT-concern | Concordant (moderate risk, widely prescribed) | High usage (~42% of FQ prescriptions) |
| Ciprofloxacin | Low-Moderate | Less-DICT-concern | Slight Overuse (global trend matches UAE) | Very high usage (~40% of FQ prescriptions) |
Comparative analysis between the UAE Department of Health real-world fluoroquinolone prescribing patterns and FDA DICTrank risk categorization for drug-induced cardiotoxicity (DICT). DICTrank classification sourced from publicly available FDA labeling datasets covering 1,318 approved and withdrawn drugs. UAE prescription trends extracted from MOHAP Open Health Data Portal (2018–2023).
Footnote: Most-DICT-concern drugs contain boxed warnings for severe QT prolongation risk. Less-DICT-concern drugs include those listing QT-related adverse events in the adverse reaction or precaution sections without boxed warnings.

### Slide 12
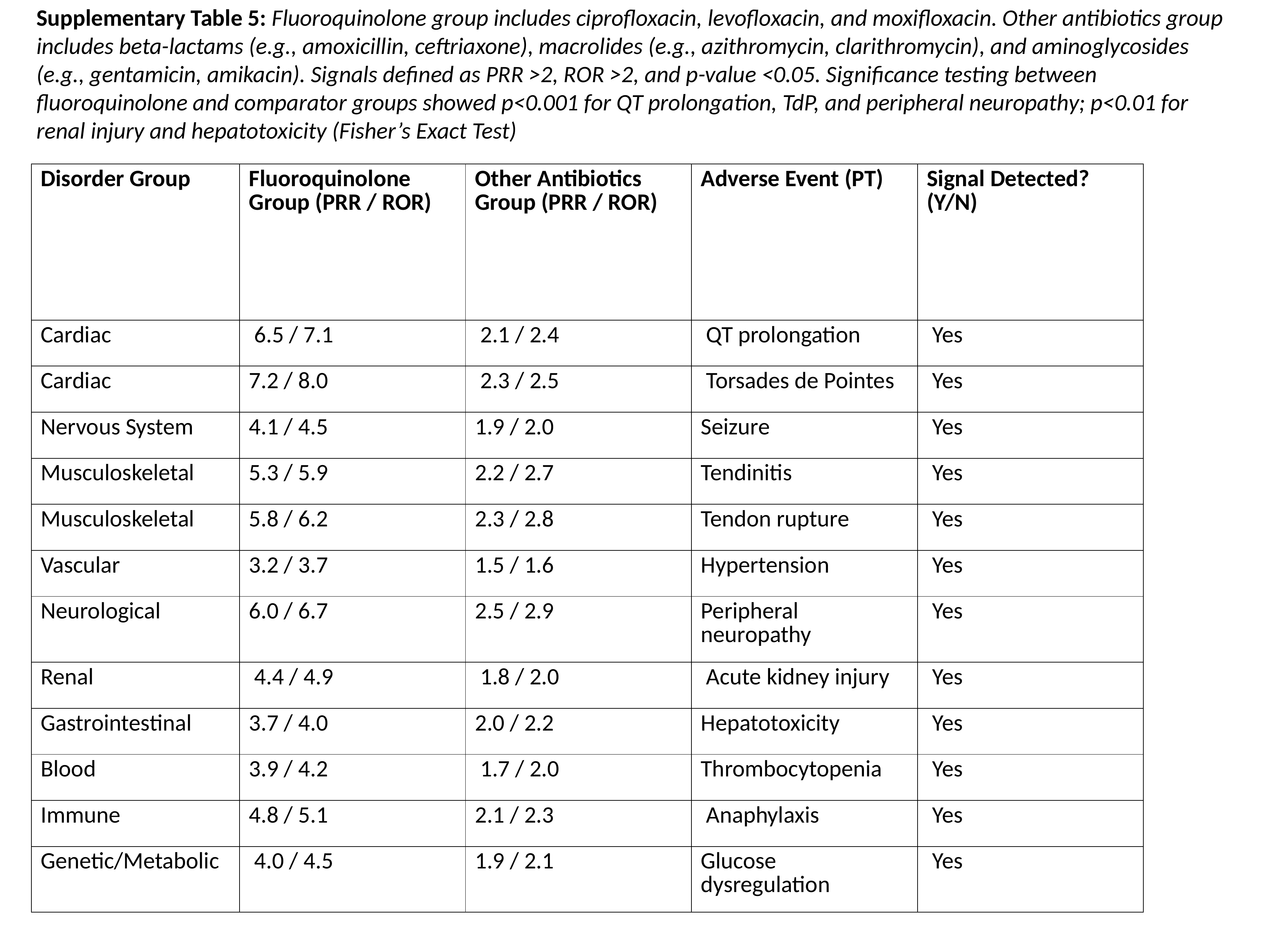

Supplementary Table 5: Fluoroquinolone group includes ciprofloxacin, levofloxacin, and moxifloxacin. Other antibiotics group includes beta-lactams (e.g., amoxicillin, ceftriaxone), macrolides (e.g., azithromycin, clarithromycin), and aminoglycosides (e.g., gentamicin, amikacin). Signals defined as PRR >2, ROR >2, and p-value <0.05. Significance testing between fluoroquinolone and comparator groups showed p<0.001 for QT prolongation, TdP, and peripheral neuropathy; p<0.01 for renal injury and hepatotoxicity (Fisher’s Exact Test)
| Disorder Group | Fluoroquinolone Group (PRR / ROR) | Other Antibiotics Group (PRR / ROR) | Adverse Event (PT) | Signal Detected? (Y/N) |
| --- | --- | --- | --- | --- |
| Cardiac | 6.5 / 7.1 | 2.1 / 2.4 | QT prolongation | Yes |
| Cardiac | 7.2 / 8.0 | 2.3 / 2.5 | Torsades de Pointes | Yes |
| Nervous System | 4.1 / 4.5 | 1.9 / 2.0 | Seizure | Yes |
| Musculoskeletal | 5.3 / 5.9 | 2.2 / 2.7 | Tendinitis | Yes |
| Musculoskeletal | 5.8 / 6.2 | 2.3 / 2.8 | Tendon rupture | Yes |
| Vascular | 3.2 / 3.7 | 1.5 / 1.6 | Hypertension | Yes |
| Neurological | 6.0 / 6.7 | 2.5 / 2.9 | Peripheral neuropathy | Yes |
| Renal | 4.4 / 4.9 | 1.8 / 2.0 | Acute kidney injury | Yes |
| Gastrointestinal | 3.7 / 4.0 | 2.0 / 2.2 | Hepatotoxicity | Yes |
| Blood | 3.9 / 4.2 | 1.7 / 2.0 | Thrombocytopenia | Yes |
| Immune | 4.8 / 5.1 | 2.1 / 2.3 | Anaphylaxis | Yes |
| Genetic/Metabolic | 4.0 / 4.5 | 1.9 / 2.1 | Glucose dysregulation | Yes |

### Slide 13
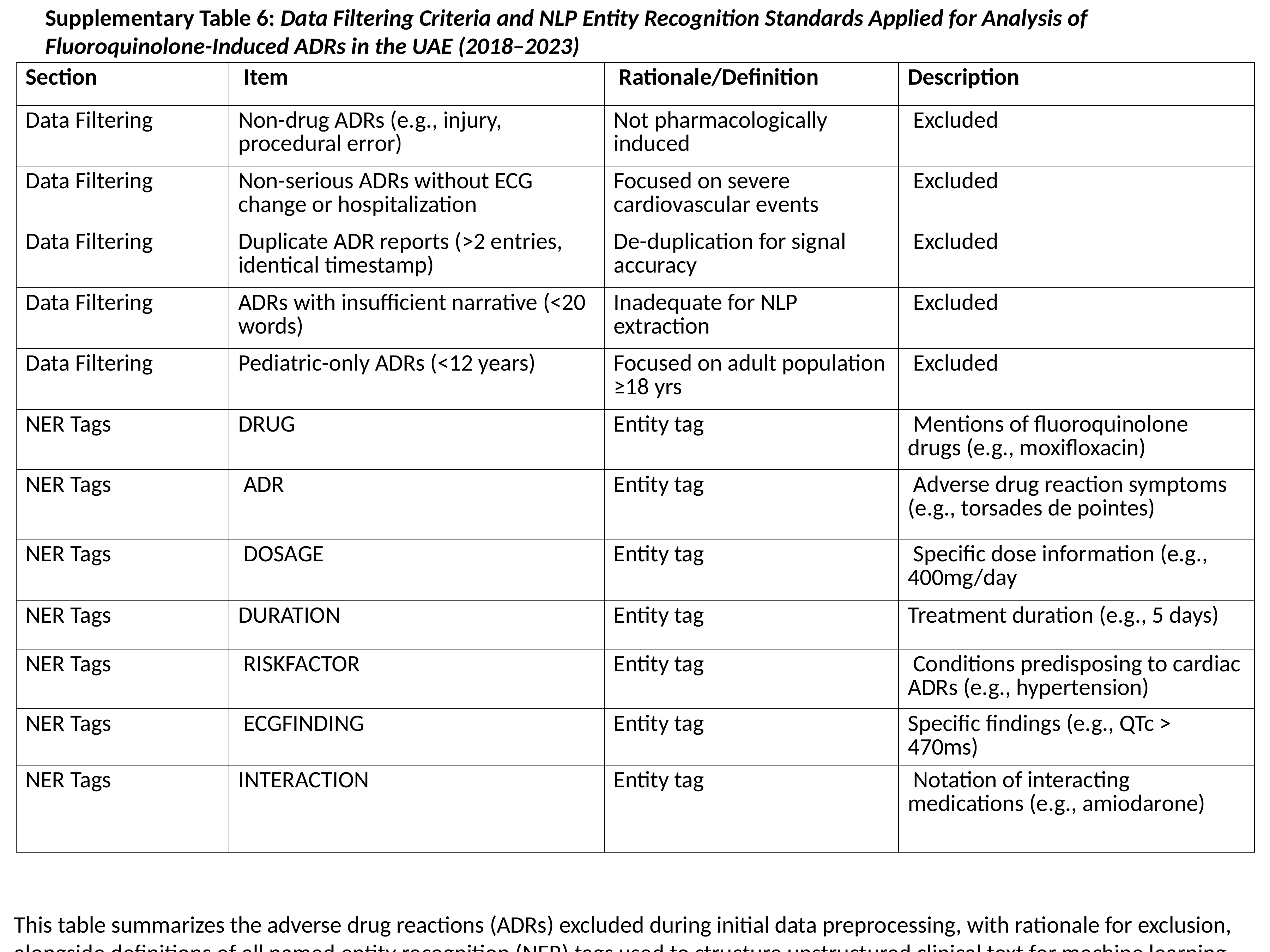

Supplementary Table 6: Data Filtering Criteria and NLP Entity Recognition Standards Applied for Analysis of Fluoroquinolone-Induced ADRs in the UAE (2018–2023)
| Section | Item | Rationale/Definition | Description |
| --- | --- | --- | --- |
| Data Filtering | Non-drug ADRs (e.g., injury, procedural error) | Not pharmacologically induced | Excluded |
| Data Filtering | Non-serious ADRs without ECG change or hospitalization | Focused on severe cardiovascular events | Excluded |
| Data Filtering | Duplicate ADR reports (>2 entries, identical timestamp) | De-duplication for signal accuracy | Excluded |
| Data Filtering | ADRs with insufficient narrative (<20 words) | Inadequate for NLP extraction | Excluded |
| Data Filtering | Pediatric-only ADRs (<12 years) | Focused on adult population ≥18 yrs | Excluded |
| NER Tags | DRUG | Entity tag | Mentions of fluoroquinolone drugs (e.g., moxifloxacin) |
| NER Tags | ADR | Entity tag | Adverse drug reaction symptoms (e.g., torsades de pointes) |
| NER Tags | DOSAGE | Entity tag | Specific dose information (e.g., 400mg/day |
| NER Tags | DURATION | Entity tag | Treatment duration (e.g., 5 days) |
| NER Tags | RISKFACTOR | Entity tag | Conditions predisposing to cardiac ADRs (e.g., hypertension) |
| NER Tags | ECGFINDING | Entity tag | Specific findings (e.g., QTc > 470ms) |
| NER Tags | INTERACTION | Entity tag | Notation of interacting medications (e.g., amiodarone) |
This table summarizes the adverse drug reactions (ADRs) excluded during initial data preprocessing, with rationale for exclusion, alongside definitions of all named entity recognition (NER) tags used to structure unstructured clinical text for machine learning analysis. Data filtered according to WHO-UMC guidelines and UAE Department of Health standards.

### Slide 14
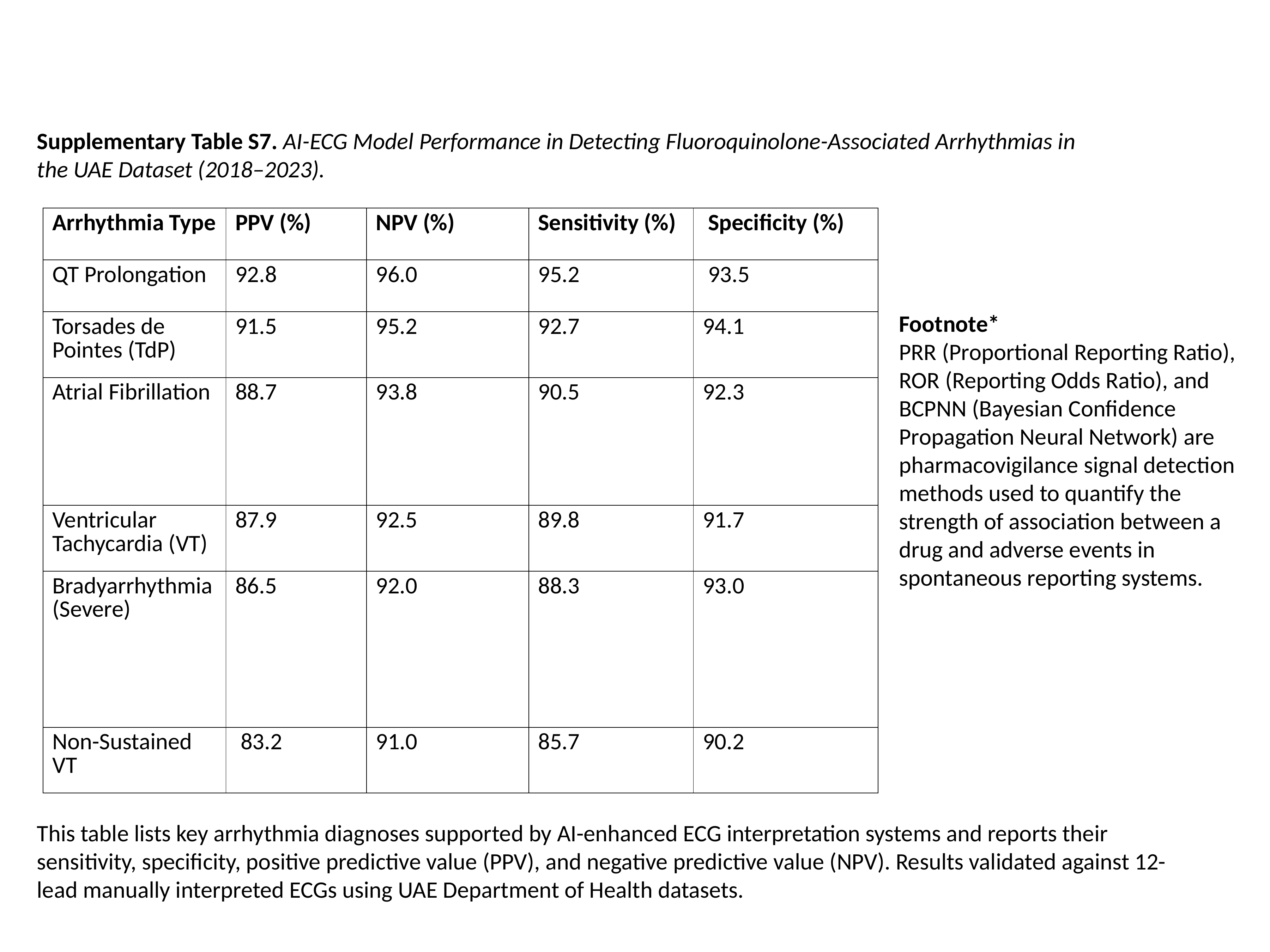

Supplementary Table S7. AI-ECG Model Performance in Detecting Fluoroquinolone-Associated Arrhythmias in the UAE Dataset (2018–2023).
| Arrhythmia Type | PPV (%) | NPV (%) | Sensitivity (%) | Specificity (%) |
| --- | --- | --- | --- | --- |
| QT Prolongation | 92.8 | 96.0 | 95.2 | 93.5 |
| Torsades de Pointes (TdP) | 91.5 | 95.2 | 92.7 | 94.1 |
| Atrial Fibrillation | 88.7 | 93.8 | 90.5 | 92.3 |
| Ventricular Tachycardia (VT) | 87.9 | 92.5 | 89.8 | 91.7 |
| Bradyarrhythmia (Severe) | 86.5 | 92.0 | 88.3 | 93.0 |
| Non-Sustained VT | 83.2 | 91.0 | 85.7 | 90.2 |
Footnote*
PRR (Proportional Reporting Ratio), ROR (Reporting Odds Ratio), and BCPNN (Bayesian Confidence Propagation Neural Network) are pharmacovigilance signal detection methods used to quantify the strength of association between a drug and adverse events in spontaneous reporting systems.
This table lists key arrhythmia diagnoses supported by AI-enhanced ECG interpretation systems and reports their sensitivity, specificity, positive predictive value (PPV), and negative predictive value (NPV). Results validated against 12-lead manually interpreted ECGs using UAE Department of Health datasets.

### Slide 15
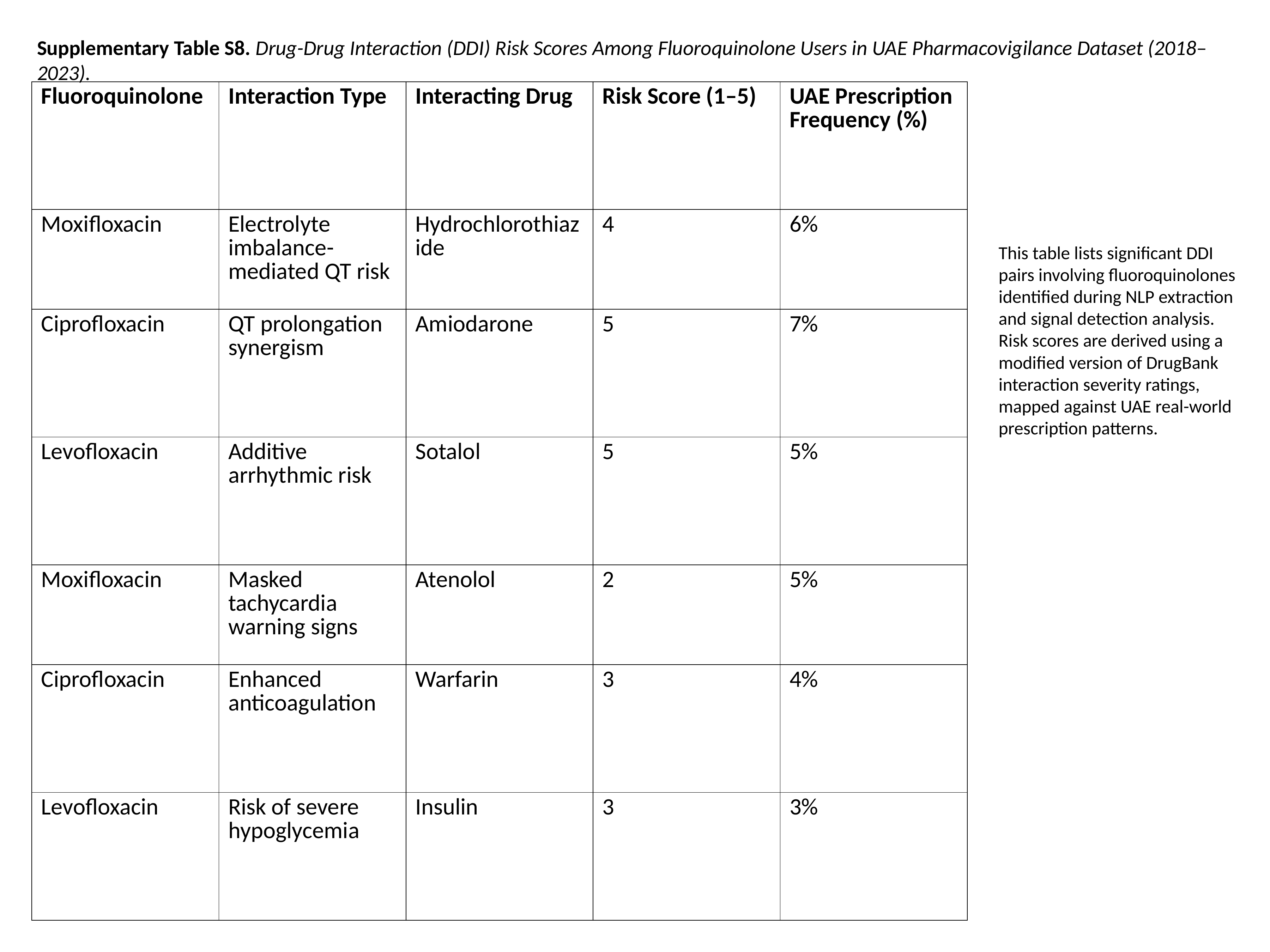

Supplementary Table S8. Drug-Drug Interaction (DDI) Risk Scores Among Fluoroquinolone Users in UAE Pharmacovigilance Dataset (2018–2023).
| Fluoroquinolone | Interaction Type | Interacting Drug | Risk Score (1–5) | UAE Prescription Frequency (%) |
| --- | --- | --- | --- | --- |
| Moxifloxacin | Electrolyte imbalance-mediated QT risk | Hydrochlorothiazide | 4 | 6% |
| Ciprofloxacin | QT prolongation synergism | Amiodarone | 5 | 7% |
| Levofloxacin | Additive arrhythmic risk | Sotalol | 5 | 5% |
| Moxifloxacin | Masked tachycardia warning signs | Atenolol | 2 | 5% |
| Ciprofloxacin | Enhanced anticoagulation | Warfarin | 3 | 4% |
| Levofloxacin | Risk of severe hypoglycemia | Insulin | 3 | 3% |
This table lists significant DDI pairs involving fluoroquinolones identified during NLP extraction and signal detection analysis. Risk scores are derived using a modified version of DrugBank interaction severity ratings, mapped against UAE real-world prescription patterns.

### Slide 16
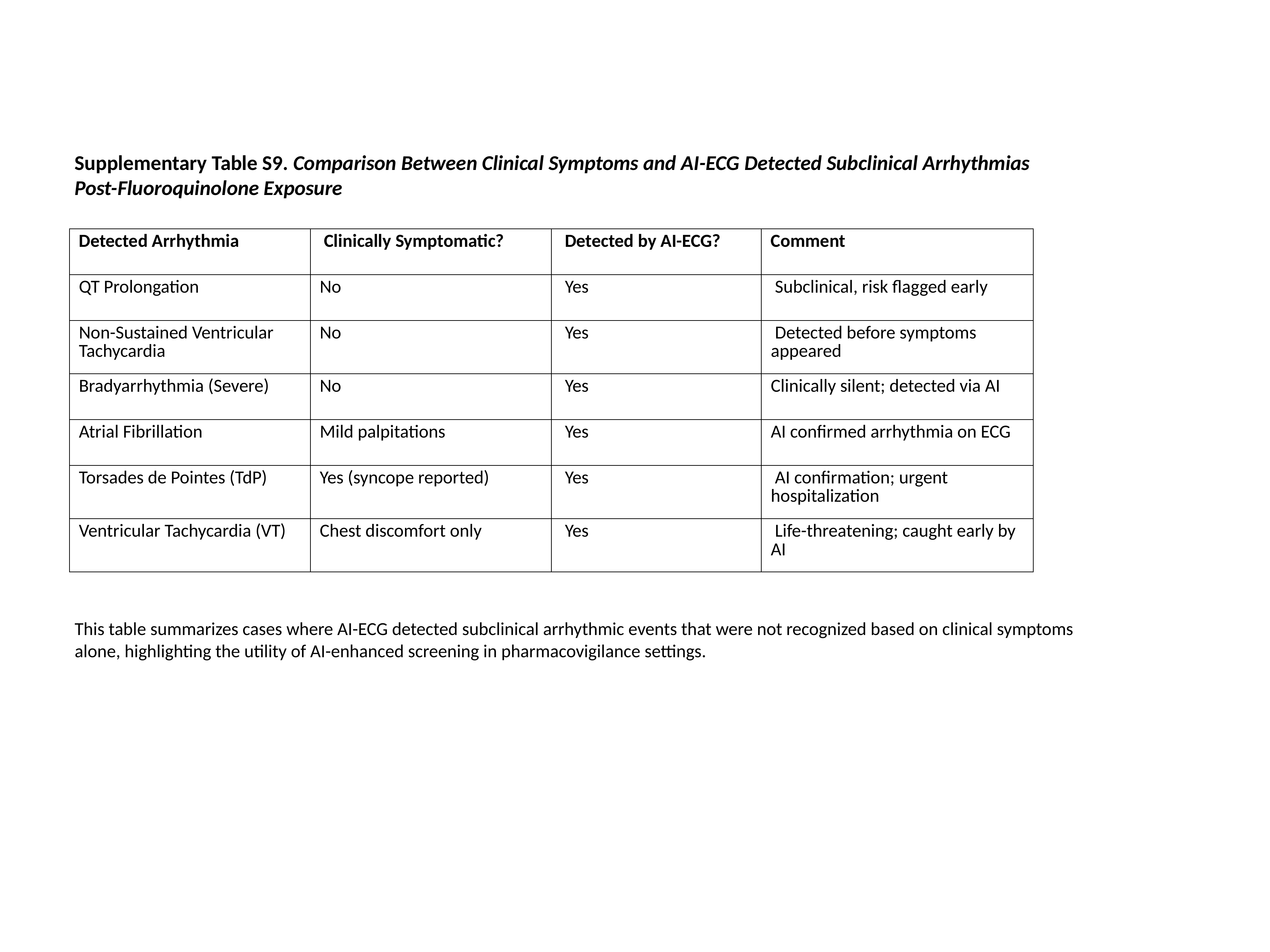

Supplementary Table S9. Comparison Between Clinical Symptoms and AI-ECG Detected Subclinical Arrhythmias Post-Fluoroquinolone Exposure
| Detected Arrhythmia | Clinically Symptomatic? | Detected by AI-ECG? | Comment |
| --- | --- | --- | --- |
| QT Prolongation | No | Yes | Subclinical, risk flagged early |
| Non-Sustained Ventricular Tachycardia | No | Yes | Detected before symptoms appeared |
| Bradyarrhythmia (Severe) | No | Yes | Clinically silent; detected via AI |
| Atrial Fibrillation | Mild palpitations | Yes | AI confirmed arrhythmia on ECG |
| Torsades de Pointes (TdP) | Yes (syncope reported) | Yes | AI confirmation; urgent hospitalization |
| Ventricular Tachycardia (VT) | Chest discomfort only | Yes | Life-threatening; caught early by AI |
This table summarizes cases where AI-ECG detected subclinical arrhythmic events that were not recognized based on clinical symptoms alone, highlighting the utility of AI-enhanced screening in pharmacovigilance settings.

### Slide 17
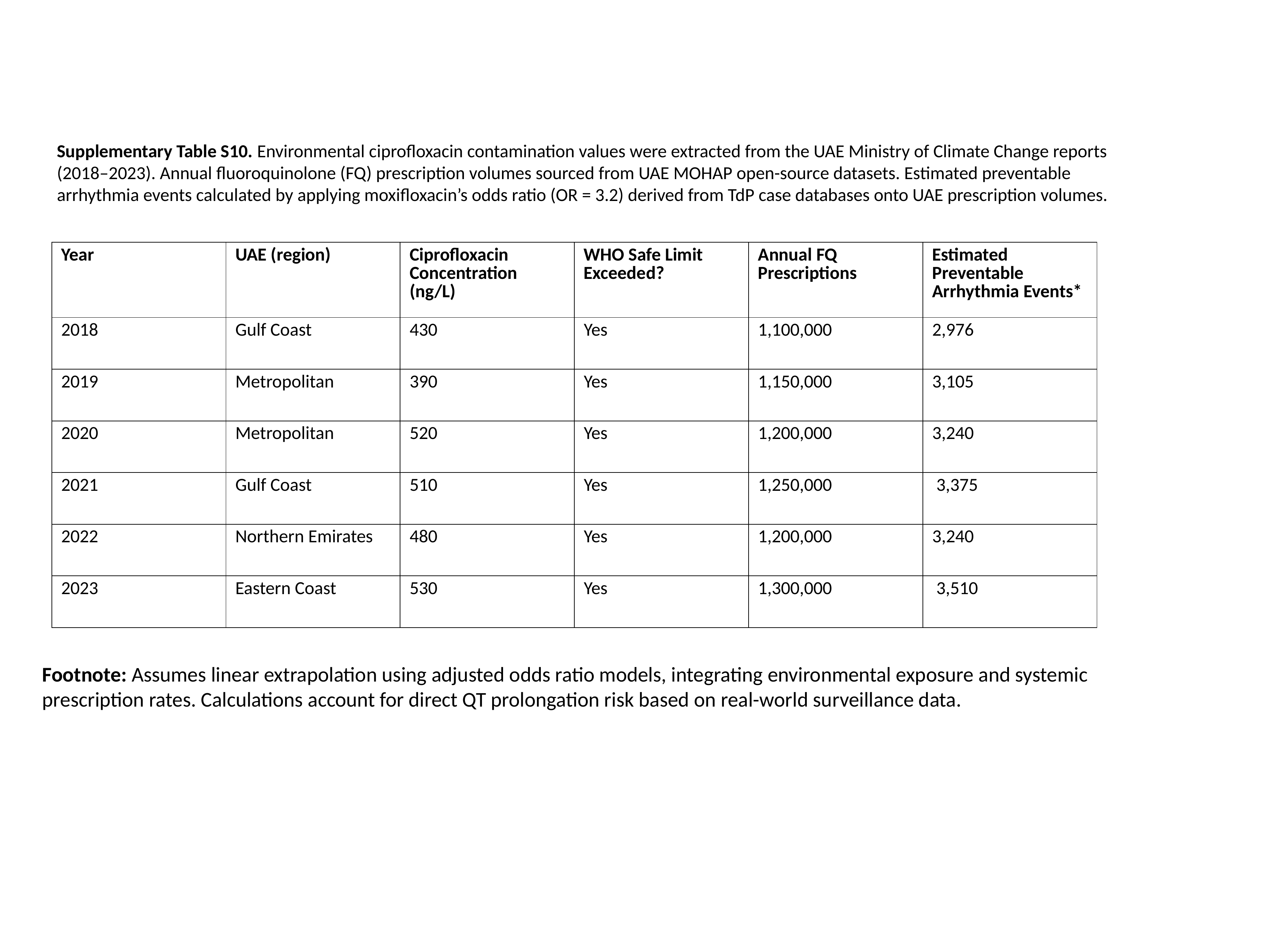

Supplementary Table S10. Environmental ciprofloxacin contamination values were extracted from the UAE Ministry of Climate Change reports (2018–2023). Annual fluoroquinolone (FQ) prescription volumes sourced from UAE MOHAP open-source datasets. Estimated preventable arrhythmia events calculated by applying moxifloxacin’s odds ratio (OR = 3.2) derived from TdP case databases onto UAE prescription volumes.
| Year | UAE (region) | Ciprofloxacin Concentration (ng/L) | WHO Safe Limit Exceeded? | Annual FQ Prescriptions | Estimated Preventable Arrhythmia Events\* |
| --- | --- | --- | --- | --- | --- |
| 2018 | Gulf Coast | 430 | Yes | 1,100,000 | 2,976 |
| 2019 | Metropolitan | 390 | Yes | 1,150,000 | 3,105 |
| 2020 | Metropolitan | 520 | Yes | 1,200,000 | 3,240 |
| 2021 | Gulf Coast | 510 | Yes | 1,250,000 | 3,375 |
| 2022 | Northern Emirates | 480 | Yes | 1,200,000 | 3,240 |
| 2023 | Eastern Coast | 530 | Yes | 1,300,000 | 3,510 |
Footnote: Assumes linear extrapolation using adjusted odds ratio models, integrating environmental exposure and systemic prescription rates. Calculations account for direct QT prolongation risk based on real-world surveillance data.
